# Limited English Proficiency and Rates of Dilated Eye Examinations Among U.S. Adults with Diabetes

**DOI:** 10.64898/2026.11.16.26368231

**Authors:** Stanley Z. Lam, Emily Xing, Grace Sun

**Affiliations:** MD Program, Weill Cornell Medicine, 1300 York Ave, New York, New York, 10021, USA; Department of Ophthalmology, Weill Cornell Medicine, 1305 York Ave, New York, New York, 10021, USA

**Keywords:** Limited English proficiency, dilated fundus examination, diabetes, diabetic retinopathy screening

## Abstract

**Purpose:** To determine whether English proficiency is associated with diabetic retinopathy screening rates among patients with diabetes in the United States.

**Design:** A retrospective, cross-sectional study.

**Participants:** Individuals with diabetes who responded to the Medicare Expenditure Panel Survey (MEPS), a nationally representative survey of U.S. households (2013-2022).

**Methods:** Survey-weighted, multivariable logistic regression.

**Main Outcome Measures:** Adjusted odds ratio (aOR) of 1) having ever received a dilated fundus examination (DFE) and 2) having received consecutive year DFEs.

**Results:** The analytic sample included 11,441 respondents representing a weighted sample of 15,107,166 U.S. civilians with diabetes per year from 2013-2022. The mean age was 62 years (SD: 14 years), and 50% were female. Individuals with limited English proficiency (LEP) comprised 11.6% of the cohort. Respondents with LEP had lower odds of ever having received a DFE compared to English-proficient respondents (aOR=0.729, 95% CI: 0.541-0.976). The association between having consecutive year DFEs and English proficiency was non-significant (aOR=1.189, 95% CI: 0.917-1.541). In sensitivity analyses using a definition of LEP as speaking English less than “well,” LEP was associated with lower odds of ever having a DFE (aOR=0.632, 95% CI: 0.460-0.869) and higher odds of consecutive year exams (aOR=1.415, 95% CI: 1.057-1.896).

**Conclusions:** LEP status was associated with lower odds of ever having had a DFE and equal or greater odds of having consecutive year DFEs. Efforts to reduce language-related disparities for patients with LEP should focus on improving initial access to ophthalmologic care.

## Introduction

Over 29 million people in the United States have a diagnosis of diabetes, and an estimated 11 million more have diabetes that is undiagnosed.^1^ Diabetic retinopathy is a microvascular complication of diabetes and the leading cause of blindness among American adults aged 20-74 years old.^2,3^ A 2021 study estimated that 9.6 million Americans have diabetic retinopathy (DR), and of those 1.8 million have had vision-threatening advanced microvascular complications.^4^ DR can remain clinically asymptomatic until advanced stages,^5^ but early treatment can reduce the risk of severe vision loss by up to 94%.^3^ Therefore, timely diagnosis and intervention are crucial for preventing vision loss from DR.

The dilated fundus examination (DFE) is an essential component in the primary prevention of DR. The American Academy of Ophthalmology (AAO) and the American Diabetes Association (ADA) both recommend annual dilated eye examinations for adult patients with diabetes,^6^ but only 18 to 60% of people with diabetes undergo the DFEs at the recommended frequency.^7^ Prior studies utilizing nationally representative datasets such as the Medicare Expenditure Panel Survey^8^ and the National Health and Nutrition Examination Survey^9^ have found that reduced adherence to recommended DR screening guidelines is associated with multiple sociodemographic factors, including lower education, not having insurance, and lower household income.^10,11^ Notably, language is one sociodemographic factor that has not been examined in these nationally representative studies as a correlate of DR screening rates.

Preferred language and English proficiency are critical social determinants of healthcare access, quality, and outcomes.^12^ There are over 25 million people in the United States with limited English proficiency (LEP).^13^ Several federal laws prohibit discrimination against patients with LEP and mandate that they receive language services such as translated documents and professional interpretation.^14^ However, in practice, such services are not always provided, and LEP status has been associated with reduced access to healthcare and poorer health outcomes.^12^ A study of 13,590 eyes of patients presenting for cataract surgery found that 18.1% of patients with LEP had diabetic retinopathy vs. 5.8% of English-proficient patients.^15^ This finding raises an important health equity concern of whether patients with LEP have adequate access to ophthalmology care.

The literature on the association between language and access to eye care has been limited to regional studies. The Los Angeles Latino Eye Study (N=5,455) found that compared to English-speakers, Spanish speakers were less likely to have an eye care visit in the past year but were equally likely to have ever had a dilated eye examination.^16^ A study of N=974 patients with diabetes in St. Louis reported that non-English speakers had a 48% greater odds of having visited an ophthalmologist in the last 2 years.^17^ To our knowledge, no study has examined the association between English proficiency and DFE adherence in a nationally representative sample.

In this study, we utilized a decade of Medicare Expenditure Panel Survey data (2013-2022) to examine the relationship between language and dilated eye examination rates in a nationally representative sample of civilians with diabetes in the United States. This study had two aims: first, to determine the association between English proficiency and ever having received dilated eye screening, and second, to determine the association between English proficiency and guideline-adherent dilated eye screening.

## Methods

### Data Source

We performed a retrospective cross-sectional analysis using survey data from the Medical Expenditure Panel Survey (MEPS) from 2013 to 2022. MEPS is sponsored by the Agency for Healthcare Research and Quality, a component of the U.S. Department of Health and Human Services, and covers topics including use of medical services, access to care, and health conditions. Each year, households in the United States that participated in the prior year’s National Health Interview Survey (National Center for Health Statistics) are invited to participate in MEPS.^18^ A stratified multistage sampling method is applied to achieve a sample that is representative of the U.S. noninstitutionalized civilian population.^11,19^ Participants are interviewed up to 5 times over a 2-year period. As fewer participants were included in each panel during the COVID-19 pandemic, participants were interviewed up to 9 times over a 4-year period.^20^ Participants aged 18 and older who answer “Yes” that a doctor or other health professional has told them that they had diabetes or sugar diabetes receive a paper copy of the Diabetes Care Survey,^21^ which is available in English and Spanish. MEPS data harmonized over the period 2013-2022 was accessed through the Integrated Public Use Microdata Series (IPUMS).^22^ MEPS data until 2022 was used because Diabetes Care Survey data for 2023 and beyond are not publicly available. Participants aged 18 and older who completed the Diabetes Care Survey component of MEPS were included, and those with unknown or missing responses to language variables (SPKOTHLANG, OTHLANGFAM, SPKENGWELL, OTHLANGWELL) and diabetic eye examination variables (DIAEYEXNV, DIAEYEXPVYR, DIAEYEXCALYR) were excluded. Because the data are de-identified and publicly accessible, the Weill Cornell Medicine IRB determined that IRB review and approval were not required (submission number: 26-06030447-01). The study was in conformity with all country, federal, or state laws and was performed in adherence to the tenets of the Declaration of Helsinki.

### Outcome Measures

The survey item pertaining to DFE frequency in the Diabetes Care Survey was as follows: “Which of the following year(s) did you have an eye exam in which your pupils were dilated? This would have made you temporarily sensitive to bright light. Mark all that apply.” Participants were able to select up to five options: during the current calendar year of the survey, during the previous calendar year, before the previous calendar year, during the subsequent calendar year, or never. For participants who took the Diabetes Care Survey multiple times, data from the most recent year was used. We used Tran et al.’s (2013) strategy for defining DFE outcome measures.^11^ The two main outcomes were ever having received a dilated eye examination and receiving consecutive year dilated eye exams, which was defined as having an eye examination in the current calendar year and previous calendar year.

### Language Measures

Participants were asked to rate their English proficiency (“Very well,” “Well,” “Not well,” “Not at all”) if they spoke a language other than English at home (2018-2022) or if they reported having someone in the family who spoke a language other than English (2013-2018). We used the official U.S. government definition of LEP—speaking English less than “very well”^23^—for the key independent variable of self-reported English proficiency (Very well vs. LEP). In sensitivity analyses, we used an alternative definition of LEP as those speaking English “Not well” or “Not at all” (Very well / Well vs. LEP). Other language measures included the type of non-English language spoken at home (“Spanish” or “Another language”).

### Other measures

Other covariates were selected based on prior studies^10,11^ and by using the Andersen Healthcare Model of Utilization as a theoretical framework for explaining DFE frequency. According to this framework, healthcare utilization can be explained by a) predisposing factors such as demographic characteristics, b) enabling factors, e.g., income and insurance, that enable people to access services, and c) need factors, which motivate people to use health services.^24^ The predisposing factors selected as covariates were age, sex (male or female), race and ethnicity (Non-Hispanic White, Non-Hispanic Black, Hispanic, Non-Hispanic Asian, and Other), and education level (no high school diploma, high school diploma, some college but no 4-year degree, Bachelor’s degree or higher). Enabling factors were household income as a percentage of the federal poverty line (FPL) (below FPL, 100–124% of FPL, 125–199% of FPL, 200–399% of FPL, and above 400% of FPL), insurance status (uninsured, Medicaid and other public insurance, private insurance), and usual source of care (no usual source of care, has a usual source of care). Need factors were self-rated health (poor, fair, good, very good, excellent) and diabetes medication use (insulin or oral medication use vs. no insulin or oral medication use).

### Statistical Analysis

To achieve prevalence estimates per year that were representative of the U.S. noninstitutionalized civilian population with diabetes over the period 2013-2022, analyses performed accounted for the Diabetic Care Survey person-level weights, clustering, and stratification design. Participants with missing values for English proficiency or eye examination outcomes were removed from analysis. We used cross-tabulations to calculate prevalence of sociodemographic factors by English proficiency. Continuous variables and categorical variables were compared via design-based t-tests and Pearson’s Χ^2^ with Rao and Scott adjustment, respectively. To account for multiple comparisons, q values were calculated from p values using a Bonferroni adjustment. Survey-adjusted bivariate logistic regressions were used to determine the association between English proficiency and the main outcomes (ever received a DFE and receiving consecutive year DFEs). Then, survey-adjusted multivariable logistic regressions were performed adjusting for all remaining covariates with the following reference levels: age, sex (reference: male), race (reference: White), education (reference: no high school diploma), income (reference: under FPL), insurance (reference: no insurance), usual source of care (reference: no usual source of care), self-rated health, and diabetes medication use (reference: none). As a post-hoc sensitivity analysis, we reran the multivariable logistic regressions with the alternative definition of LEP as those speaking English “Not well” or “Not at all.” We also explored whether the type of non-English language spoken at home (Spanish or Non-English Non-Spanish) was an effect modifier for the relationship between English proficiency and the two outcomes through stratification analyses. Adjusted odds ratios (aOR) and 95% confidence intervals (95% CI) for each correlate were reported. Two-sided p < 0.05 was considered statistically significant. Collinearity across variables was assessed using variance inflation factors (VIFs), with VIF > 5 representing the presence of collinearity. All statistical analysis was performed using R (4.5.2).^25^ R packages used included survey,^26^ ipumsr,^27^ and forester.^28^

## Results

### Sample characteristics

The unweighted analytic sample included 11,441 respondents, representing a weighted sample size of 15,107,166 U.S. civilians with diabetes on average per year over the period 2013 to 2022. An average of 13,349,168 (88.4%) individuals were proficient in English and 1,757,998 (11.6%) had limited English proficiency. The mean age of the analytic sample was 62 years (SD: 14 years), and 50% were female. Table 1 shows differences in sociodemographic characteristics between LEP and English-proficient participants with diabetes. Compared to those who were proficient in English, participants with LEP were more likely to be female (54% vs. 49%, q=0.018), have lower education attainment (q<0.001), not have a usual source of care (16% vs. 8.3%, q<0.001), and have poorer self-rated health (q<0.001). Moreover, a higher percentage of participants with LEP were Hispanic (69% vs 9.5%) or Asian (22% vs 3.7%), and more participants with LEP were uninsured (12% vs 2.7%) or used public health insurance (54% vs 37%). Overall, 11% of respondents had never had a DFE, and 62% of respondents with diabetes in the sample did not have consecutive year dilated eye exams. Fewer LEP participants had ever received a DFE in the past (79% vs. 90%, q<0.001). Similarly, fewer LEP vs. English-proficient respondents had DFEs in consecutive years (31% vs. 39%, q=0.001).

**Table 1.** Sociodemographic and health characteristics of U.S. adults with diabetes by English proficiency from 2013 to 2022. Values shown as weighted percentages of the total analytic cohort (unweighted N=11,441). For age, a design-based t-test was used. For all other measures, Pearson’s X2: Rao and Scott adjustment were performed. Bonferroni adjustment was used to obtain q-values from p-values.

| Characteristic | **Overall**<br>Weighted N = 15,187,168 | **English proficient**<br>Weighted N = 13,416,587 | **Limited English proficient**<br>Weighted N = 1,770,581 | **q-value** |
|---|---|---|---|---|
| Age, years (mean; SD) | 62 (14) | 62 (14) | 61 (13) | >0.9 |
| Female sex | 50% | 49% | 54% | 0.018 |
| Race and ethnicity |  |  |  | <0.001 |
| White | 59% | 66% | 5.2% |  |
| Black | 16% | 17% | 3.0% |  |
| Hispanic | 16% | 9.5% | 69% |  |
| Asian | 5.9% | 3.7% | 22% |  |
| Other | 3.5% | 3.8% | 0.7% |  |
| Education level |  |  |  | <0.001 |
| No HS diploma | 17% | 12% | 50% |  |
| HS diploma | 33% | 35% | 24% |  |
| Some college, no 4-year degree | 18% | 20% | 6.8% |  |
| Bachelor's degree or higher | 31% | 33% | 17% |  |
| Unknown | 0.6% | 0.5% | 1.7% |  |
| Household income as percentage of federal poverty line (FPL) |  |  |  | <0.001 |
| Below FPL | 14% | 13% | 24% |  |
| 100-124% of FPL | 5.6% | 5.3% | 7.7% |  |
| 125-199% of FPL | 16% | 15% | 22% |  |
| 200-399% of FPL | 29% | 30% | 28% |  |
| Above 400% of FPL | 35% | 37% | 18% |  |
| Insurance status |  |  |  | <0.001 |
| Uninsured | 3.8% | 2.7% | 12% |  |
| Medicaid and other public | 39% | 37% | 54% |  |
| Private | 57% | 60% | 34% |  |
| Has a usual source of care |  |  |  | <0.001 |
| No usual source of care | 9.2% | 8.3% | 16% |  |
| Yes, has a usual source of care | 90% | 91% | 83% |  |
| Unknown | 0.9% | 0.8% | 1.3% |  |
| Self-rated health |  |  |  | <0.001 |
| Poor | 25% | 24% | 35% |  |
| Fair | 6.2% | 6.2% | 6.2% |  |
| Good | 41% | 41% | 39% |  |
| Very Good | 21% | 22% | 14% |  |
| Excellent | 3.4% | 3.4% | 3.0% |  |
| Unknown | 3.6% | 3.6% | 3.2% |  |
| On diabetes medication |  |  |  | >0.9 |
| No | 13% | 13% | 14% |  |
| Yes | 87% | 87% | 86% |  |
| Unknown | <0.1% | <0.1% | <0.1% |  |
| Ever had an eye exam | 89% | 90% | 79% | <0.001 |
| Had consecutive year eye exams | 38% | 39% | 31% | 0.001 |

### Dilated fundus exams by English proficiency

In bivariate analyses, LEP status was associated with lower odds of ever having had a DFE (OR=0.416, 95% CI: 0.340-0.508) and receiving consecutive year DFEs (OR=0.697, 95% CI: 0.581-0.836). After adjusting for predisposing, enabling, and need characteristics in survey-weighted multivariable logistic regressions, respondents with LEP had a 27.1% lower odds of ever having received a DFE compared to English-proficient respondents (aOR=0.729, 95% CI: 0.541-0.976; Table 2, Figure S1) and the association between having consecutive year DFEs and English proficiency was no longer significant (aOR=1.189, 95% CI: 0.917-1.541; Table 2, Figure S2).

**Table 2.** The association between limited English proficiency status with ever receiving dilated eye exams and receiving consecutive year dilated eye exams. DFE = dilated fundus examination, LEP = limited English proficiency, CI = confidence interval.

| Model | Adjusted odds ratio | 95% CI | p value |
|---|---|---|---|
| **Main analysis** – entire cohort<br>LEP = speaking English "Well," "Not well" or "Not at all" |  |  |  |
| Ever had DFE ~ LEP + Covariates |  |  |  |
| LEP | 0.728 | 0.541 to 0.979 | <0.05 |
| Consecutive year DFEs ~ LEP + Covariates |  |  |  |
| LEP | 1.187 | 0.917 to 1.536 | 0.193 |
| **Sensitivity analysis** – entire cohort<br>LEP (alternative definition) = speaking English "Not well" or "Not at all" |  |  |  |
| Ever had DFE ~ LEP + Covariates |  |  |  |
| LEP | 0.632 | 0.460 to 0.869 | <0.01 |
| Consecutive year DFEs ~ LEP + Covariates |  |  |  |
| LEP | 1.415 | 1.057 to 1.896 | <0.05 |

In a sensitivity analysis with LEP defined as speaking English “Not well” / “Not at all”, LEP status was associated with a 36.8% lower odds of having ever received a DFE (aOR=0.632, 95% CI: 0.460-0.869) but a 45.1% greater odds of receiving consecutive year DFEs (aOR=1.415, 95% CI: 1.057-1.896; Table 2). As an exploratory analysis, we stratified the sample into a Spanish-speaking subcohort (unweighted N=2,237) and Non-English Non-Spanish-speaking subcohort (unweighted N=719). In the Spanish-speaking subcohort, the aORs for ever having received a DFE was 0.806 (95% CI: 0.541-1.200) and for receiving consecutive year DFEs was 1.115 (95% CI: 0.794-1.566). In the Non-English Non-Spanish-speaking subcohort, the aORs for ever having received a DFE was 0.696 (95% CI: 0.349-1.387) and for receiving consecutive year DFEs was 0.803 (95% CI: 0.474-1.360).

## Discussion

In this nationally representative study of civilians with diabetes in the United States, we investigated the relationship between limited English proficiency status and adherence to dilated eye examination. Our findings of overall rates of ever receiving a DFE and consecutive year dilated eye exams over the period 2013 to 2022 are consistent with Tran et al.’s analysis of the MEPS sample with diabetes from 2002-2013, which found that 10% had never had a DFE and 60% did not have consecutive year dilated eye exams.^11^ This suggests that adherence to dilated eye exams has not measurably improved across the past two decades for patients with diabetes in the United States. Notably, Eppley et al. (2019) used a different nationally representative survey database—the National Health and Nutrition Examination Survey—and likewise concluded that guideline-adherent screening rates did not improve at a population level between the period 2005-2006 and 2015-2016.^10^

Prior analyses of sociodemographic factors associated with DFE rates among patients with diabetes spanning multiple U.S. states have not examined language as a correlate of DFE^10,11,29^ or had a limited number of non-English speaking respondents.^30^ This study found that after adjusting for other sociodemographic variables, limited English proficiency status was independently associated with a lower odds of ever having received a DFE but not associated with likelihood of consecutive year DFEs. Importantly, when we used a more stringent definition of LEP as those speaking English less than “Well” in sensitivity analyses, patients with LEP had lower odds of ever having had a DFE but greater odds of adherence to guideline-recommended dilated eye screening. These findings, while seemingly contradictory, pinpoint the key disparity for patients with LEP in the context of DFE adherence. Once LEP patients with diabetes have had their first DFE, they are not less likely to adhere to guideline-recommended DFE rates—in fact, those with particularly limited English proficiency may be adhering to recommended screening at rates higher than that of English-proficient patients. Thus, interventions seeking to reduce language-based disparities in dilated eye examination rates should target patients who have never received specialized ophthalmology care. Lastly, our analyses stratified by type of language spoken at home revealed no significant associations between LEP and either outcome. While the non-significance may be due to reduced power from smaller sample sizes, the findings suggest that language is likely not an effect modifier for the relationship between LEP status and DFE rates.

Patients with LEP may be presenting with worse outcomes due to delays in receiving necessary screening and care. Gill et al. reported that LEP patients presenting for cataract surgery had a 100% greater odds of diabetic retinopathy compared to English-proficient patients after adjusting for sociodemographic variables. Moreover, they found that patients with LEP had worse mean best-corrected visual acuity at presentation and higher rates of macular edema.^15^ Internationally, a study in Singapore similarly found associations between non-English preference and increased DR rates in an English-dominant population.^31^ In a qualitative study of Spanish-speakers in the Michigan Screening and Intervention for Glaucoma and Eye Health through Telemedicine Program (MI-SIGHT) trial, language barriers were cited as a primary obstacle to accessing eye care, with participants citing difficulties filling medical forms in English and lack of staff who understand their language as factors discouraging them from seeking eye care.^32^ Another qualitative study of linguistic minority groups in Canada specifically identified language as a barrier to attending diabetic retinopathy screening.^33^ Therefore, we hypothesize that language barriers experienced by LEP patients with diabetes reduces initial engagement in dilated eye screening, which in turn contributes to higher rates of diabetic retinopathy in this population.

To improve initial uptake of DR screening for patients with LEP, we suggest the following clinic-based and community-based approaches. First, facilitating language-concordant interactions at the stage of initial patient contact with clinics may help reduce barriers to accessing needed ophthalmologic care. An intervention that could improve LEP patients’ initial access to ophthalmologic care at the clinic level is the implementation of language-concordant automated telephone systems. Linares et al. (2024) called 124 ambulatory clinics across Boston, Massachusetts, and found that non-English language phone recordings were used by only 22% of specialty clinics, compared to 52% of primary care clinics.^34^ Their follow-up qualitative study found that language-discordant phone systems contributed to reluctance to call specialty clinics and delays in care.^35^ In an abstract by Valdivia Camacho et al. (2025), using an English-only automated phone scheduling system was found to result in a reduced appointment completion rate for LEP vs. English proficient patients.^36^ Other language interventions that have been shown to improve access to specialty care for non-English speaking patients include having bilingual staff and appointment reminders in the patient’s preferred language.^37^ In one study of Spanish- and Arabic-speakers with LEP, 47% of Spanish-speakers and 54% of Arabic-speakers reported difficulty understanding their appointment reminders.^38^ One potential strategy to address this communication barrier is the use of artificial intelligence (AI)-based communication tools, such as multi-lingual chatbots to send reminders and instructions in patients’ preferred languages.^39^

Second, free community eye screenings not only provide eye exams for patients who have never had them but also can connect patients with specialty eye care.^40^ In the MI-SIGHT trial, participants from low-income communities received comprehensive eye disease screening; 7% percent of patients with diabetes screened reported never having had a dilated eye examination.^41^ As our study found that LEP patients are more than twice as likely to never have had a dilated eye examination, community screening programs with bilingual staff could provide an avenue through which many LEP patients are connected to specialized eye care for the first time. Another strategy may be to increase AI diabetic retinopathy screening technology in the primary care setting, which has been shown to increase the likelihood of receiving subsequent ophthalmologic care.^42^

Lastly, strengthening the referral pathway to ophthalmologists via patient navigators may increase screening rates for LEP patients who have never received eye care. Patient navigators can assist patients with scheduling appointments, appointment reminders, and transportation assistance. In the New York City Screening and Intervention for Glaucoma and Eye Health through Telemedicine Program (NYC-SIGHT) trial, patients who were referred for an in-office eye exam were less likely to attend their initial visit if they were Spanish-speaking. However, patients randomized to the Navigator Intervention group had a significantly higher odds of adherence. Patient navigators are a promising intervention for enhancing initial access to care for patients with LEP.^43^

### Limitations

A key strength of this study using the MEPS database is achieving estimates and associations that are nationally representative of the U.S. civilian population with diabetes. However, an important limitation of the study is that the quality of the data is limited by the accuracy of patient self-report. While patient self-report of a diabetes diagnosis has been shown to have high positive and negative predictive value compared to data extracted from the medical record,^44^ other aspects, such as DFE frequency, may be subject to underreporting. To investigate the possibility that self-report of DFEs could be subject to recall bias, we examined the two most recent years of survey data for participants who took the Diabetes Care Survey multiple times. At a per-participant level, we found that up to 34% of responses regarding DFE frequency were not concordant between survey years (e.g., reporting that they had a DFE in 2017, and then reporting in 2018 that they did not have a DFE in 2017). We dealt with this limitation by including only the most recent year of survey data available for all participants. Another limitation is that while the Diabetes Care Survey is available in English and Spanish, LEP participants who prefer to use other languages such as Mandarin or Vietnamese would have to rely on other sources such as family members to help them complete the survey. This barrier may make those who prefer to speak non-English non-Spanish languages less likely to complete the survey, which would result in their underrepresentation in the cohort. Future studies should examine the association of English proficiency with DFE frequency within specific language subcohorts, to see if language barriers affect certain subgroups disproportionately.

## Conclusion

In conclusion, this study found that LEP status is associated with a reduced odds of ever having received an exam, but rates of consecutive year exams are similar or higher among patients with LEP compared to English proficient patients. To our knowledge, this study is the first to demonstrate an association between English proficiency and DFE frequency among a nationally representative sample of patients with diabetes in the United States. This study sheds light into a key disparity for diabetic patients with LEP: initial access to DFE screening. Clinic based-interventions (e.g., language-concordant automated phone systems and appointment reminders) and community-based programs (e.g., patient navigator and screening programs) may facilitate first-time connections between LEP patients and specialized ophthalmologic care, improving health equity for patients with LEP.

## Data Availability

All data analyzed in the study can be accessed at: https://meps.ipums.org/meps/

## Acknowledgements

a. Funding/Support: This research did not receive any specific grant from funding agencies in the public, commercial, or not-for-profit sectors.
b. Financial Disclosures: The authors have no financial conflicts of interests to disclose relevant to this manuscript.
c. Other Acknowledgments: The authors would like to thank Dr. Paul J. Christos (Weill Cornell Medicine Associate Research Professor) for his expert advice on the statistical analysis for this project.

## Author contributions

Stanley Z. Lam: Conceptualization, Data Curation, Formal Analysis, Investigation, Methodology, Software, Visualization, Writing – Original Draft Preparation, Writing – Review & Editing

Emily Xing: Formal Analysis, Investigation, Software, Writing – Original Draft Preparation, Writing – Review & Editing

Grace Sun: Investigation, Methodology, Supervision, Writing – Original Draft Preparation, Writing – Review & Editing

## Declaration of generative AI and AI-assisted technologies in the manuscript preparation process

Statement: ChatGPT (OpenAI), Claude (Anthropic) and OpenEvidence were used (June to September 2026) to: refine the analytic plan for the project, identify literature sources, and to troubleshoot R code for analysis. Claude was used to check for grammar for select portions of the manuscript; however, none of the writing, figures, or tables in this manuscript were generated by AI. The authors collectively agreed on the analysis strategy, reviewed analytic models with a biostatistician, and reviewed all literature sources. S.L. and G.S. had full access to all the data in the study and take responsibility for the integrity of the data and the accuracy of the data analysis.

