## Supplemental Figures 1 and 2 for "Limited English Proficiency and Rates of Dilated Eye Examinations Among U.S. Adults with Diabetes"

This Supplementary File contains forest plots showing effect sizes for all covariates in the main analyses.

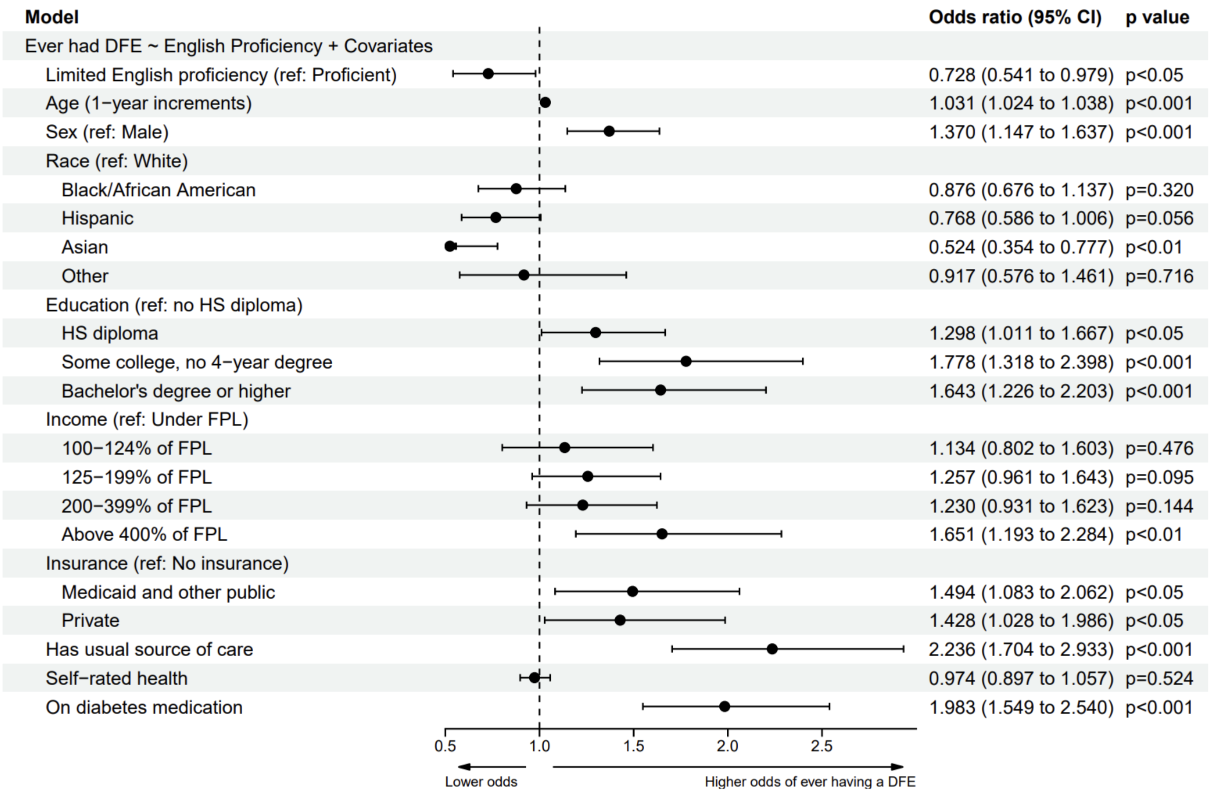


**Supplementary Figure 1.** Multivariable logistic regression for the association between ever receiving a dilated examination and English proficiency. Limited English proficiency is defined as speaking English “Well,” “Not well,” or “Not at all.” The black circles on the forest plot represent the point estimate of the adjusted odds ratios for each correlate, and the horizontal bars represent the 95% confidence intervals (CI). DFE = dilated fundus examination, HS = high school, FPL = federal poverty level, ref = reference level.

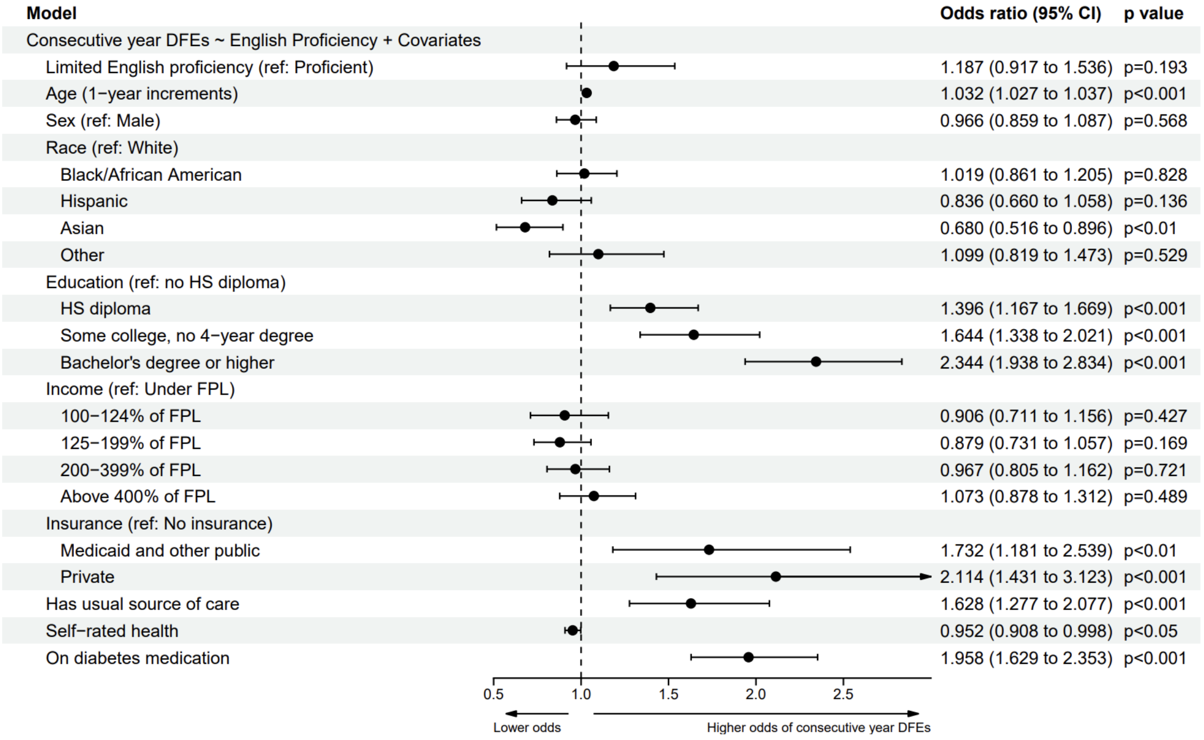


**Supplementary Figure 2.** Multivariable logistic regression for the association between consecutive year dilated eye examinations and English proficiency. Limited English proficiency is defined as speaking English "Well," "Not well," or "Not at all." The black circles on the forest plot represent the point estimate of the adjusted odds ratios for each correlate, and the horizontal bars represent the 95% confidence intervals (CI). DFE = dilated fundus examination, HS = high school, FPL = federal poverty level, ref = reference level.
